# Changes in outpatient care patterns and subsequent outcomes during the COVID-19 pandemic: A retrospective cohort analysis from a single payer healthcare system

**DOI:** 10.1101/2022.03.07.22272032

**Authors:** Finlay A. McAlister, Zoe Hsu, Yuan Dong, Carl van Walraven, Jeffrey A. Bakal

**Affiliations:** The Division of General Internal Medicine, Faculty of Medicine and Dentistry, University of Alberta, Edmonton, Alberta, Canada; The Alberta Strategy for Patient Oriented Research Support Unit, Edmonton, Canada; The Division of General Internal Medicine, Ottawa Hospital and the Ottawa Health Research Institute, Ottawa, Canada; Provincial Research Data Services, Alberta Health Services, Canada

## Abstract

**Background:** There have been rapid shifts in outpatient care models during the COVID-19 pandemic but the impact of these changes on patient outcomes are uncertain. We designed this study to examine ambulatory outpatient visit patterns and outcomes between March 1, 2019 to February 29, 2020 (pre-pandemic) and from March 1, 2020 to February 28, 2021 (pandemic).

**Methods:** We conducted a population-based retrospective cohort study of all 3.8 million adults in the Canadian province of Alberta, which has a single payer healthcare system, using linked administrative data. We examined all outpatient physician encounters (virtual or in-person) and outcomes (emergency department visits, hospitalizations, or deaths) in the next 30- and 90-days.

**Results:** Although in-person outpatient visits declined by 38.9% in the year after March 1, 2020 (10,142,184 vs. 16,592,599), the increase in virtual visits (7,152,147; 41.4% of total) meant that total outpatient encounters increased by 4.1% in the first year of the pandemic. Outpatient care and prescribing patterns remained stable for adults with ambulatory-care sensitive conditions (ACSC): 97.2% saw a primary care physician (median 6 visits), 59.0% had at least one specialist visit, and 98.5% were prescribed medications (median 9) in the year prior to the pandemic compared to 96.6% (median 3 in-person and 2 virtual visits), 62.6%, and 98.6% (median 8 medications) during the first year of the pandemic. In the first year of the pandemic, virtual outpatient visits were associated with less subsequent healthcare encounters than in-person ambulatory visits, particularly for patients with ACSC (9.2% vs. 10.4%, aOR 0.89 [95% confidence interval 0.87-0.92] at 30 days and 26.9% vs. 29.3%, aOR 0.93 [0.92-0.95] at 90 days).

**Conclusions:** The shifts in outpatient care patterns caused by the COVID-19 pandemic did not disrupt prescribing or follow-up for patients with ACSC and did not worsen post-visit outcomes.

**Funding:** None

**Registration:** None

**KEY MESSAGES:** *What is already known on this topic:* There have been rapid shifts in outpatient care models during the COVID-19 pandemic but outcomes are uncertain.

*What this study adds:* Total outpatient encounters increased by 4% in the first year of the pandemic due to a rapid increase in virtual visits (which made up 41% of all outpatient encounters). Prescribing patterns and frequency of follow-up were similar in the first year after onset of the pandemic in adults with ambulatory-care sensitive conditions. Compared to in-person visits, virtual outpatient visits were associated with less subsequent healthcare encounters, particularly for patients with ambulatory-care sensitive conditions (11% less at 30 days and 7% less at 90 days).

*How this study might affect research, practice or policy:* Our data provides reassurance that the shifts in outpatient care patterns caused by the COVID-19 pandemic did not negatively impact follow-up, prescribing, or outcomes for patients with ACSC. Further research is needed to define which patients and which conditions are most suitable for virtual outpatient visits and, as with all outpatient care, the optimal frequency of such visits.

## INTRODUCTION

Due to the COVID-19 pandemic, there has been a substantial shift in outpatient medical care from in-person office visits to virtual care (mostly via telephone).[1-5] While several studies reported that patients with chronic conditions were less likely to be seen *by any modality* (even virtually) after the onset of the pandemic,[6-10] these studies only examined the first few months after pandemic onset and the impact of these changes in ambulatory treatment patterns on patient outcomes is unclear. It is entirely possible that the introduction of virtual physician assessments would eventually increase the frequency of outpatient care given the decreased barriers for both patients (i.e. decreased travel time, decreased costs) and physicians (i.e. possible decreased time per patient, no travel required to attend clinic). However, the lack of in-person contact and the absence of information from physically examining patients could also negatively influence patient outcomes.

In this study, we examined changes in ambulatory healthcare patterns after the onset of the COVID-19 pandemic in Alberta, Canada. First, we wanted to see whether the volume of virtual and in-person outpatient physician assessments changed during this time. Second, we explored whether outcomes for ambulatory care sensitive conditions (ACSC, defined by the Canadian Institute of Health Information as asthma, chronic obstructive pulmonary disease, heart failure, coronary disease, hypertension, diabetes, and epilepsy) were different after virtual or in-person outpatient visits.

## METHODS

### Study Design

This was a retrospective cohort study. We included all adult Albertans using physician services in two sequential time periods: March 1 2019 to February 29 2020 (classified as ‘pre-pandemic’); and March 1, 2020 to February 28, 2021 (pandemic). Our study ended at this time because of data availability.

### Data Sources and Study Sample

This study linked population-based health administrative datasets in Alberta, Canada for 3.8 million adults. All health care in the province is publicly funded with universal access and without user fees at the point of care.

Several datasets were linked deterministically via encrypted unique health identifier number to create our study’s analytical dataset. The *Discharge Abstract Database* (DAD) captures all acute care hospitalizations recording admission and discharge date, as well as up to 25 diagnoses and 16 procedures indicated with ICD-10 and CCI codes, respectively. The *Ambulatory Care Database* (ACD) captures all Emergency Department assessments and hospital-based physician office visits recording the date along with up to 10 diagnostic codes. The *Healthcare Provider Claims Database* (HPCD) captures claims for all physician visits (including those shadow-billed by salaried physicians) recording the date and up to 3 diagnoses. The *Pharmacy Information Network* (PIN) captures the type, dates, and doses of all medication dispensations from community pharmacies. The *Alberta Health Care Insurance Registry* captures all patient demographics and includes addresses. The comprehensiveness of the databases we used in this study have been previously established.[11,12]

### Identifying and Classifying Physician Visits

We retrieved all encounters with physicians, emergency departments (ED), or hospitals during the 2 sequential study time periods. All physician encounters were classified as ambulatory outpatient, ED, or hospital based. We limited physician assessments to one type per patient per day.

Alberta Health modified the Alberta Schedule of Medical Benefits physician billing codes on March 17, 2020 to include new codes for virtual (telephone or video) visits; prior to this, physicians were required – with few exceptions - to see patients in person to qualify for billing. We classified all outpatient visits during the study as virtual (denoted by the codes 03.01AD, 03.01CC, 03.03CV, 03.03FV, 03.08CV, 08.19CX, 08.19CV, and 08.19CW) or in-person (denoted by the codes 03.03A, 03.03AZ, 03.03C, 03.03F, 03.03FA, 03.03FZ, 03.04A, 03.04AZ, 03.04J, 03.07A, 03.07AZ, 03.07B, 03.08A, 03.08I, 03.08AZ, 03.08IZ, 08.19C, or 08.19G).

To compare outcomes after virtual vs. in-person visits, our primary analysis defined outpatient visit type based on the first visit for each patient in each of the time periods (pre-pandemic and during the pandemic). In a sensitivity analysis, we explored the robustness of our findings by restricting the outcome analyses to only those patients with a single outpatient visit in the outcome period.

### Outcomes

We examined the proportion of patient visits followed by an ED visit, a hospitalization, death, or a composite of any of the 3 events within 30 days and 90 days of their index outpatient visit. Patients who presented to an ED and were subsequently admitted to hospital would have contributed an event to both the estimate of ED visits and the estimate of hospitalizations. The composite outcome was calculated in two ways: first, as a sum of events (i.e., total number of ED visits, hospitalizations and deaths) to capture overall utilization (our primary analysis) and second, as a binary outcome to indicate if any of the three outcomes (ED visit, hospitalization, or death) had occurred (sensitivity analysis).

### Comorbidities

We used ICD-9 and ICD-10-CA case definitions previously validated in Alberta for any hospitalizations, any ED visits, and any outpatient visits in the 2 years prior to and including the index visit to identify comorbidities for each patient.[11] We used previously validated case definitions from CIHI to identify those patients with at least one ACSC (eAppendix Table 1: asthma, chronic obstructive pulmonary disease, heart failure, coronary disease, hypertension, diabetes, or epilepsy).

### Statistical Analysis

We report care patterns by patient demographics and subsequent outcomes by presence/absence of ACSC. Differences were assessed for statistical significance (p<0.05) using the Chi-square test or the Mann–Whitney U test / Wilcoxon rank-sum test as appropriate. To calculate adjusted odds ratios for each of the outcomes after outpatient visits, we included age, sex, Pampalon deprivation index, and the Charlson comorbidity score. For the composite outcome, we used logistic regression models to analyze the sum of events composite (our primary analysis) and used zero-inflated Poisson models for the binary composite outcome (since the zero-inflated Poisson is used in cases where a large percentage of the count variable outcomes are zero). All statisical analyses were done using SAS v.9.4 (Cary, N.C.) and figures were generated using R 4.1.2.

### Ethics

The University of Alberta Health Research Ethics Board approved this study (Pro00086861) and granted a waiver for individual patient consent as the investigators were only provided with de-identified data after linkage to conform with provincial privacy regulations.

## RESULTS

The number of adults who had at least one healthcare encounter declined from 2,807,604 in 2019-20 to 2,684,694 in 2020-21 (Tables 1 and 2). Between 2019-20 and the first year of the pandemic (Tables 1 and 2), the proportion of community-dwelling Albertan adults presenting to an ED at least once decreased (from 40.1% to 34.3% for those with ACSC and from 25.5% to 22.3% for those without ACSC), as did the proportion requiring hospitalization (from 16.2% to 14.8% of those with ACSC and from 5.5% to 5.3% of those without ACSC). However, the proportion of long-term care residents presenting to an ED (74.7% vs. 73.5%) or being hospitalized (66.4% vs. 67.6%) remained relatively stable.

**Table 1:** Health service use by adults in Alberta in the year prior to the COVID-19 pandemic.

|  | Community Dwelling Adults |  |  |  |  |  |  |  |  |  | Long Term<br>Care<br>Residents |
| --- | --- | --- | --- | --- | --- | --- | --- | --- | --- | --- | --- |
|  | Overall | Urban | Rural | 18-40 y | 40-65 y | ≥65 y | Male | Female | No ACSC | ≥1 ACSC |  |
| N | 2954024 | 2564063 | 389961 | 1151660 | 1254983 | 547381 | 1387746 | 1566278 | 2350147 | 603877 | 7047 |
| Median age<br>(IQR) | 46<br>(33, 60) | 45<br>(32, 60) | 50<br>(34, 63) | 30<br>(24, 35) | 52<br>(46, 58) | 72<br>(68, 79) | 46<br>(33, 61) | 45<br>(32, 60) | 41<br>(30, 56) | 62<br>(50, 72) | 84<br>(75, 89) |
| Female % | 1566278<br>(53.0) | 1362819<br>(53.2) | 203459<br>(52.2) | 629602<br>(54.7) | 644726<br>(51.4) | 291950<br>(53.3) | 0 (0.0) | 1566278<br>(100.0) | 0 (0, 0) | 1 (0, 1) | 4015 (57.0) |
| Median<br>Charlson Score | 0 (0, 0) | 0 (0, 0) | 0 (0, 0) | 0 (0, 0) | 0 (0, 0) | 0 (0, 1) | 0 (0, 0) | 0 (0, 0) | 0 (0, 0) | 1 (0, 1) | 2 (1, 3) |
| Urban<br>Residence | 2564063<br>(86.8) | 2564063<br>(100.0) | 0 (0.0) | 1017591<br>(88.4) | 1088919<br>(86.8) | 457553<br>(83.6) | 1201244<br>(86.6) | 1362819<br>(87.0) | 2052646<br>(87.3) | 511417<br>(84.7) | 5790 (82.2) |
| **Pampalon Material Deprivation Index** |  |  |  |  |  |  |  |  |  |  |  |
| 1 (least<br>deprived) | 546441<br>(18.5) | 536393<br>(20.9) | 10048<br>(2.6) | 216264<br>(18.8) | 232334<br>(18.5) | 97843<br>(17.9) | 255336<br>(18.4) | 291105<br>(18.6) | 455469<br>(19.4) | 90972<br>(15.1) | 1106 (15.7) |
| 2 | 538731<br>(18.2) | 503422<br>(19.6) | 35309<br>(9.1) | 210258<br>(18.3) | 234126<br>(18.7) | 94347<br>(17.2) | 249812<br>(18.0) | 288919<br>(18.4) | 438852<br>(18.7) | 99879<br>(16.5) | 1044 (14.8) |
| 3 | 548557<br>(18.6) | 491829<br>(19.2) | 56728<br>(14.5) | 214200<br>(18.6) | 236892<br>(18.9) | 97465<br>(17.8) | 256068<br>(18.5) | 292489<br>(18.7) | 438395<br>(18.7) | 110162<br>(18.2) | 1121 (15.9) |
| 4 | 585810<br>(19.8) | 474324<br>(18.5) | 111486<br>(28.6) | 221530<br>(19.2) | 251897<br>(20.1) | 112383<br>(20.5) | 276664<br>(19.9) | 309146<br>(19.7) | 458538<br>(19.5) | 127272<br>(21.1) | 1306 (18.5) |

|  | Community Dwelling Adults |  |  |  |  |  |  |  |  |  | Long Term |
| --- | --- | --- | --- | --- | --- | --- | --- | --- | --- | --- | --- |
| 5 (most deprived) | 599525 (20.3) | 457307 (17.8) | 142218 (36.5) | 243239 (21.1) | 249751 (19.9) | 106535 (19.5) | 287248 (20.7) | 312277 (19.9) | 458421 (19.5) | 141104 (23.4) | 1443 (20.5) |
| Saw a Primary Care Physician in office setting | 2733529 (92.5) | 2386790 (93.1) | 346739 (88.9) | 1045538 (90.8) | 1177364 (93.8) | 510627 (93.3) | 1254446 (90.4) | 1479083 (94.4) | 2146521 (91.3) | 587008 (97.2) | 4623 (65.6) |
| Median number of PCP office visits | 3 (2, 6) | 3 (2, 6) | 3 (1, 5) | 3 (1, 5) | 4 (2, 6) | 5 (2, 8) | 3 (1, 5) | 4 (2, 7) | 3 (1, 5) | 6 (4, 9) | 1 (0, 4) |
| Virtual visit with primary care physician | 0 (0.0) | 0 (0.0) | 0 (0.0) | 0 (0.0) | 0 (0.0) | 0 (0.0) | 0 (0.0) | 0 (0.0) | 0 (0.0) | 0 (0.0) | 0 (0.0) |
| Median number of PCP virtual visits | 0 (0.0) | 0 (0.0) | 0 (0.0) | 0 (0.0) | 0 (0.0) | 0 (0.0) | 0 (0.0) | 0 (0.0) | 0 (0.0) | 0 (0.0) | 0 (0.0) |
| Saw any specialist in office setting | 1167635 (39.5) | 1029632 (40.2) | 138003 (35.4) | 316648 (27.5) | 517840 (41.3) | 333147 (60.9) | 505958 (36.5) | 661677 (42.2) | 811311 (34.5) | 356324 (59.0) | 3432 (48.7) |
| Median number of specialist office visits | 0 (0, 1) | 0 (0, 1) | 0 (0, 1) | 0 (0, 1) | 0 (0, 1) | 1 (0, 3) | 0 (0, 1) | 0 (0, 1) | 0 (0, 1) | 1 (0, 3) | 0 (0, 2) |
| Virtual visit with specialist | 0 (0.0) | 0 (0.0) | 0 (0.0) | 0 (0.0) | 0 (0.0) | 0 (0.0) | 0 (0.0) | 0 (0.0) | 0 (0.0) | 0 (0.0) | 0 (0.0) |
| Median number of specialist virtual visits | 0 (0.0) | 0 (0.0) | 0 (0.0) | 0 (0.0) | 0 (0.0) | 0 (0.0) | 0 (0.0) | 0 (0.0) | 0 (0.0) | 0 (0.0) | 0 (0.0) |
| Had at least one medication dispensation | 2525365 (85.5) | 2187185 (85.3) | 338180 (86.7) | 907358 (78.8) | 1096088 (87.3) | 521919 (95.3) | 1136421 (81.9) | 1388944 (88.7) | 1930374 (82.1) | 594991 (98.5) | 6630 (94.1) |
| Median number of medications dispensed | 4 (1, 8) | 4 (1, 8) | 4 (2, 9) | 2 (1, 5) | 4 (2, 8) | 8 (4, 13) | 3 (1, 7) | 4 (2, 8) | 3 (1, 6) | 9 (5, 14) | 18 (11, 27) |
| Had at least one ED visit | 841137 (28.5) | 669718 (26.1) | 171419 (44.0) | 346549 (30.1) | 318362 (25.4) | 176226 (32.2) | 398123 (28.7) | 443014 (28.3) | 599029 (25.5) | 242108 (40.1) | 5265 (74.7) |
| Median number of ED visits | 0 (0, 1) | 0 (0, 1) | 0 (0, 1) | 0 (0, 1) | 0 (0, 1) | 0 (0, 1) | 0 (0, 1) | 0 (0, 1) | 0 (0, 1) | 0 (0, 1) | 1 (0, 3) |
| Had at least one hospitalization | 227059 (7.7) | 188594 (7.4) | 38465 (9.9) | 81456 (7.1) | 68975 (5.5) | 76628 (14.0) | 87796 (6.3) | 139263 (8.9) | 129308 (5.5) | 97751 (16.2) | 4681 (66.4) |
| Median number of hospitalizations | 0 (0, 0) | 0 (0, 0) | 0 (0, 0) | 0 (0, 0) | 0 (0, 0) | 0 (0, 0) | 0 (0, 0) | 0 (0, 0) | 0 (0, 0) | 0 (0, 0) | 1 (0, 2) |

Data reported as counts and percentages. ACSC ambulatory care sensitive condition, ED emergency department, IQR interquartile range.

**Table 2:** Health services use by adults in Alberta in the first year of the COVID-19 pandemic.

|  | Community Dwelling Adults |  |  |  |  |  |  |  |  |  | Long Term<br>Care<br>Residents |
| --- | --- | --- | --- | --- | --- | --- | --- | --- | --- | --- | --- |
|  | Overall | Urban | Rural | 18-40 y | 40-65 y | ≥65 y | Male | Female | No ACSC | ≥1 ACSC |  |
| N | 2834768 | 2462677 | 372091 | 1059983 | 1209696 | 565089 | 1313656 | 1521112 | 2260155 | 574613 | 5806 |
| Median age<br>(IQR) | 47<br>(33, 61) | 46<br>(33, 61) | 51<br>(34, 64) | 30<br>(24, 35) | 52<br>(46, 58) | 72<br>(68, 79) | 48<br>(34, 62) | 46<br>(33, 61) | 42<br>(31, 57) | 62<br>(51, 72) | 84<br>(75, 89) |
| Female % | 1521112<br>(53.66) | 1325216<br>(53.81) | 195896<br>(52.65) | 593657<br>(56.01) | 626702<br>(51.81) | 300753<br>(53.22) | 0 (0.00) | 1521112<br>(100.00) | 1245966<br>(55.13) | 275146<br>(47.88) | 3252<br>(56.01) |
| Median<br>number of<br>comorbidities<br>(IQR)<br>Charlson | 0 (0, 0) | 0 (0, 0) | 0 (0, 1) | 0 (0, 0) | 0 (0, 0) | 0 (0, 1) | 0 (0, 0) | 0 (0, 0) | 0 (0, 0) | 1 (0, 1) | 2 (1, 3) |
| Urban<br>Residence | 2462677<br>(86.87) | 2462677<br>(100.00) | 0 (0.00) | 937583<br>(88.45) | 1051852<br>(86.95) | 473242<br>(83.75) | 1137461<br>(86.59) | 1325216<br>(87.12) | 1972893<br>(87.29) | 489784<br>(85.24) | 4836<br>(83.29) |
| **Pampalon Material Deprivation Index** |  |  |  |  |  |  |  |  |  |  |  |
| 1 (least<br>deprived) | 523237<br>(18.46) | 513832<br>(20.86) | 9405<br>(2.53) | 197536<br>(18.64) | 224073<br>(18.52) | 101628<br>(17.98) | 241348<br>(18.37) | 281889<br>(18.53) | 434792<br>(19.24) | 88445<br>(15.39) | 949 (16.35) |
| 2 | 518679<br>(18.30) | 484766<br>(19.68) | 33913<br>(9.11) | 193841<br>(18.29) | 226718<br>(18.74) | 98120<br>(17.36) | 237117<br>(18.05) | 281562<br>(18.51) | 422262<br>(18.68) | 96417<br>(16.78) | 868 (14.95) |
| 3 | 526090<br>(18.56) | 471773<br>(19.16) | 54317<br>(14.60) | 196944<br>(18.58) | 228391<br>(18.88) | 100755<br>(17.83) | 241752<br>(18.40) | 284338<br>(18.69) | 421453<br>(18.65) | 104637<br>(18.21) | 950 (16.36) |
| 4 | 558647<br>(19.71) | 452148<br>(18.36) | 106499<br>(28.62) | 202524<br>(19.11) | 240787<br>(19.90) | 115336<br>(20.41) | 260512<br>(19.83) | 298135<br>(19.60) | 438901<br>(19.42) | 119746<br>(20.84) | 1120<br>(19.29) |

|  | Community Dwelling Adults |  |  |  |  |  |  |  |  |  | Long Term<br>Care |
| --- | --- | --- | --- | --- | --- | --- | --- | --- | --- | --- | --- |
| 5 (most<br>deprived) | 571539<br>(20.16) | 435685<br>(17.69) | 135854<br>(36.51) | 222814<br>(21.02) | 239454<br>(19.79) | 109271<br>(19.34) | 270792<br>(20.61) | 300747<br>(19.77) | 439013<br>(19.42) | 132526<br>(23.06) | 1052<br>(18.12) |
| Saw a Primary<br>Care Physician<br>in office<br>setting | 2297134<br>(81.03) | 2006818<br>(81.49) | 290316<br>(78.02) | 843262<br>(79.55) | 992273<br>(82.03) | 461599<br>(81.69) | 1033637<br>(78.68) | 1263497<br>(83.06) | 1785902<br>(79.02) | 511232<br>(88.97) | 2814<br>(48.47) |
| Median<br>number of<br>PCP office<br>visits | 2 (1, 4) | 2 (1, 4) | 2 (1, 3) | 2 (1, 3) | 2 (1, 4) | 2 (1, 4) | 2 (1, 3) | 2 (1, 4) | 2 (1, 3) | 3 (1, 6) | 0 (0, 2) |
| Virtual visit<br>with primary<br>care physician | 1703908<br>(60.11) | 1494046<br>(60.67) | 209862<br>(56.40) | 557421<br>(52.59) | 744386<br>(61.53) | 402101<br>(71.16) | 701833<br>(53.43) | 1002075<br>(65.88) | 1260359<br>(55.76) | 443549<br>(77.19) | 3678<br>(63.35) |
| Median<br>number of<br>PCP virtual<br>visits | 1 (0, 3) | 1 (0, 3) | 1 (0, 2) | 1 (0, 2) | 1 (0, 3) | 2 (0, 4) | 1 (0, 2) | 1 (0, 3) | 1 (0, 2) | 2 (1, 5) | 1 (0, 5) |
| Saw any<br>specialist in<br>office setting | 911269<br>(32.15) | 807755<br>(32.80) | 103514<br>(27.82) | 241438<br>(22.78) | 395076<br>(32.66) | 274755<br>(48.62) | 386274<br>(29.40) | 524995<br>(34.51) | 637704<br>(28.22) | 273565<br>(47.61) | 2064<br>(35.55) |
| Median<br>number of<br>specialist<br>office visits | 0 (0, 1) | 0 (0, 1) | 0 (0, 1) | 0 (0, 0) | 0 (0, 1) | 0 (0, 2) | 0 (0, 1) | 0 (0, 1) | 0 (0, 1) | 0 (0, 2) | 0 (0, 1) |
| Virtual visit<br>with specialist | 590689<br>(20.84) | 521426<br>(21.17) | 69263<br>(18.61) | 163865<br>(15.46) | 255708<br>(21.14) | 171116<br>(30.28) | 264128<br>(20.11) | 326561<br>(21.47) | 396096<br>(17.53) | 194593<br>(33.87) | 1343<br>(23.13) |
| Median<br>number of<br>specialist<br>virtual visits | 0 (0, 0) | 0 (0, 0) | 0 (0, 0) | 0 (0, 0) | 0 (0, 0) | 0 (0, 1) | 0 (0, 0) | 0 (0, 0) | 0 (0, 0) | 0 (0, 1) | 0 (0, 0) |
| Had at least one medication dispensation | 2393089<br>(84.42) | 2072510<br>(84.16) | 320579<br>(86.16) | 811131<br>(76.52) | 1044202<br>(86.32) | 537756<br>(95.16) | 1061783<br>(80.83) | 1331306<br>(87.52) | 1826683<br>(80.82) | 566406<br>(98.57) | 5386<br>(92.77) |
| Median number of medications dispensed | 3 (1, 7) | 3 (1, 7) | 4 (1, 9) | 2 (1, 4) | 4 (1, 7) | 7 (4, 12) | 3 (1, 7) | 4 (1, 8) | 3 (1, 6) | 8 (5, 13) | 17 (10, 25) |
| Had at least one ED visit | 701685<br>(24.75) | 561954<br>(22.82) | 139731<br>(37.55) | 283196<br>(26.72) | 266169<br>(22.00) | 152320<br>(26.96) | 334599<br>(25.47) | 367086<br>(24.13) | 504460<br>(22.32) | 197225<br>(34.32) | 4267<br>(73.49) |
| Median number of ED visits | 0 (0, 0) | 0 (0, 0) | 0 (0, 1) | 0 (0, 1) | 0 (0, 0) | 0 (0, 1) | 0 (0, 1) | 0 (0, 0) | 0 (0, 0) | 0 (0, 1) | 1 (0, 2) |
| Had at least one hospitalization | 205805<br>(7.26) | 170908<br>(6.94) | 34897<br>(9.38) | 75777<br>(7.15) | 61320<br>(5.07) | 68708<br>(12.16) | 80852<br>(6.15) | 124953<br>(8.21) | 120605<br>(5.34) | 85200<br>(14.83) | 3923<br>(67.57) |
| Median number of hospitalizations | 0 (0, 0) | 0 (0, 0) | 0 (0, 0) | 0 (0, 0) | 0 (0, 0) | 0 (0, 0) | 0 (0, 0) | 0 (0, 0) | 0 (0, 0) | 0 (0, 0) | 1 (0, 2) |

Data reported as counts and percentages. ACSC ambulatory care sensitive condition, ED emergency department, IQR interquartile range.

While in-person outpatient physician visits declined by 38.9% in the year after pandemic onset (from 16,592,599 to 10,142,184), the increase in virtual visits (7,152,147 during the pandemic) meant that total outpatient encounters actually increased by 4.1% in the first year of pandemic. Overall, 41.4% of outpatient visits in the first year of the COVID-10 pandemic were virtual (Figure 1). Total outpatient care patterns for community-dwelling adults with ACSC remained stable. In 2019-20, 97.2% saw a primary care physician at least once (median 6 visits) and 59.0% had at least one specialist visit, compared to 96.6% and 62.6% during the first year of the pandemic. Moreover, 98.5% received at least one prescription in 2019-20 and the median number of prescriptions dispensed was 9, while in the first year of the pandemic 98.6% received at least one prescription and the median number of prescriptions was 8. Care patterns and prescriptions for long-term care residents also stayed stable after pandemic onset.

**Figure 1:**
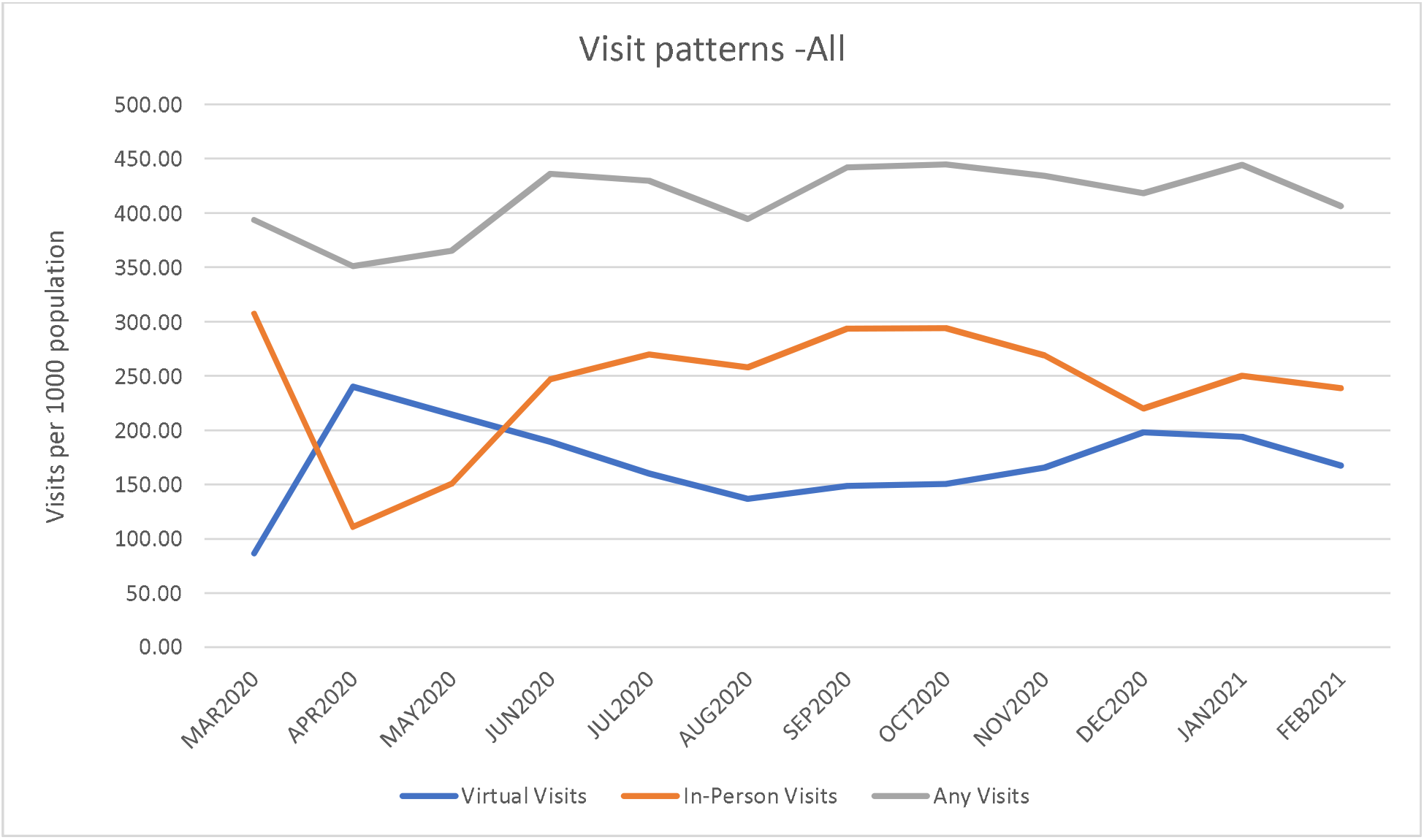
Monthly outpatient visits during the first year of the COVID-19 pandemic, per thousand adults.

Although absolute rates were higher in patients with ACSC or after specialist visits (Table 3), outcomes within 30 days of an outpatient physician visit exhibited similar changes pre/post pandemic onset in both those with (eFigure 1) or without (eFigure 2) ACSC. For example, ED visits within 30 days of an outpatient visit decreased during the first year of the pandemic compared to 2019-20 in both groups (by 25.7% in patients with ACSC and 19.0% in those without ACSC), as did hospitalizations (by 15.3% and 3.0%). Although there was an increase in mortality between 2019-20 and 2020-21, the absolute increases were small: 0.2 extra deaths per thousand outpatient visits within 30 days in both those with and those without ACSC.

**Table 3:** Outcomes after outpatient visits (reported per 1000 visits) in the year prior to the pandemic and the first year of the pandemic.

|  | In the subsequent 30 days |  |  |  | In the subsequent 90 days |  |  |  |
| --- | --- | --- | --- | --- | --- | --- | --- | --- |
|  | ED visit | Hospitalization | Death | Any Event | ED visit | Hospitalization | Death | Any Event |
| Year prior to the Pandemic |  |  |  |  |  |  |  |  |
| **Patients with ACSC** |  |  |  |  |  |  |  |  |
| PCP Visit | 100.0 | 22.3 | 0.8 | 123.1 | 268.4 | 61.3 | 3.3 | 333.0 |
| Specialist visit | 112.9 | 45.4 | 1.9 | 160.2 | 326.2 | 123.6 | 7.5 | 457.3 |
| Any physician visit | 102.2 | 26.2 | 1.0 | 129.4 | 278.2 | 71.9 | 4.1 | 354.2 |
| **Patients without ACSC** |  |  |  |  |  |  |  |  |
| PCP Visit | 50.8 | 5.7 | 0.2 | 56.7 | 125.7 | 15.1 | 0.9 | 141.7 |
| Specialist visit | 45.8 | 13.5 | 0.5 | 59.8 | 129.3 | 34.1 | 1.8 | 165.2 |
| Any physician visit | 50.1 | 6.7 | 0.3 | 57.1 | 126.2 | 17.5 | 1.0 | 144.7 |
| First year of the Pandemic |  |  |  |  |  |  |  |  |
| **Patients with ACSC** |  |  |  |  |  |  |  |  |
| PCP in-person visit | 79.8 | 19.1 | 0.6 | 99.5 | 220.8 | 54.1 | 3.3 | 278.2 |
| PCP virtual visit | 67.6 | 20.0 | 1.9 | 89.5 | 194.4 | 56.9 | 5.8 | 257.1 |
| Any PCP visit | 75.2 | 19.4 | 1.1 | 95.7 | 210.9 | 55.2 | 4.3 | 270.3 |
| Specialist in-person visit | 82.3 | 40.1 | 2.2 | 124.7 | 252.5 | 105.4 | 8.1 | 366.1 |
| Specialist virtual visit | 74.0 | 30.2 | 1.7 | 105.9 | 243.4 | 91.0 | 6.0 | 340.5 |
| Any specialist visit | 79.5 | 36.8 | 2.1 | 118.4 | 249.5 | 100.6 | 7.4 | 357.5 |
| Any physician in-person visit | 80.2 | 22.7 | 0.9 | 103.8 | 226.2 | 62.9 | 4.1 | 293.2 |
| Any physician virtual visit | 68.6 | 21.5 | 1.9 | 91.9 | 201.6 | 61.9 | 5.9 | 269.4 |
| Any physician visit | 75.9 | 22.2 | 1.2 | 99.4 | 217.1 | 62.5 | 4.8 | 284.4 |
| **Patients without ACSC** |  |  |  |  |  |  |  |  |
| PCP in-person visit | 43.3 | 5.2 | 0.2 | 48.8 | 108.9 | 14.1 | 1.0 | 123.9 |
| PCP virtual visit | 37.1 | 6.3 | 0.9 | 44.3 | 98.3 | 17.8 | 2.3 | 118.3 |
| Any PCP visit | 41.4 | 5.5 | 0.5 | 47.3 | 105.5 | 15.2 | 1.4 | 122.1 |
| Specialist in-person visit | 36.6 | 11.6 | 0.6 | 48.8 | 101.8 | 27.9 | 2.0 | 131.7 |
| Specialist virtual visit | 34.0 | 13.1 | 0.4 | 47.5 | 109.1 | 36.1 | 1.8 | 146.9 |
| Any specialist visit | 35.6 | 12.2 | 0.5 | 48.3 | 104.5 | 30.9 | 1.9 | 137.4 |
| Any physician in-person visit | 42.5 | 6.0 | 0.3 | 48.8 | 108.0 | 15.8 | 1.1 | 124.9 |
| Any physician virtual visit | 36.6 | 7.4 | 0.8 | 44.8 | 100.0 | 20.7 | 2.2 | 122.9 |
| Any physician visit | 40.6 | 6.5 | 0.5 | 47.5 | 105.4 | 17.4 | 1.5 | 124.2 |

During the first year of the pandemic, outcomes after virtual visits differed from those after in-person visits (Tables 3 and 4, eFigures 3 and 4). For example, amongst patients with ACSC, virtual outpatient visits (compared to in-person visits) were associated with less subsequent emergency department visits (6.9% vs. 8.0%, adjusted odds ratios [aOR] 0.91 [95% CI 0.89-0.94]), slightly less hospitalizations (2.2% vs. 2.3%, aOR 0.93 [0.89-0.96]), but slightly more deaths (0.19% vs. 0.09%, aOR 1.87 [1.61-2.17]) in the next 30 days. Even extending followup to 90 days, virtual visits were still associated with less emergency department visits (20.2% vs. 22.6%, aOR 0.94 [95% CI 0.92-0.95]), slightly less hospitalizations (6.2% vs. 6.3%, aOR 0.96 [95% CI 0.94-0.99]), but slightly more deaths (0.59% vs. 0.41%, aOR 1.25 [95%CI 1.15-1.35]) compared to outcomes after in-person visits. As a result, total composite events were less after virtual outpatient visits compared to in-person visits: 9.2% vs. 10.4% and aOR 0.89 [95% confidence interval 0.87-0.92] at 30 days and 26.9% vs. 29.3% with aOR 0.93 [0.92-0.95] at 90 days.

**Table 4:** Associations between virtual or in-person outpatient visits and subsequent outcomes during the first year of the COVID-19 pandemic.

|  | Outcomes in next<br>30 days for patients<br>with ACSC | Outcomes in next<br>90 days for patients<br>with ACSC | Outcomes in next<br>30 days for<br>patients without<br>ACSC | Outcomes in next<br>90 days for<br>patients without<br>ACSC |
| --- | --- | --- | --- | --- |
| **Primary Analysis (type of visit defined by index visit)** |  |  |  |  |
| ED | 0.91 (0.89, 0.94) | 0.94 (0.92, 0.95) | 0.91 (0.89, 0.93) | 0.98 (0.97, 0.99) |
| Hospitalization | 0.93 (0.89, 0.96) | 0.96 (0.94, 0.99) | 1.19 (1.15, 1.24) | 1.30 (1.27, 1.33) |
| Death | 1.87 (1.61, 2.17) | 1.25 (1.15, 1.35) | 2.25 (1.98, 2.56) | 1.47 (1.37, 1.58) |
| Composite | 0.89 (0.87, 0.92) | 0.93 (0.92, 0.95) | 0.96 (0.94, 0.97) | 1.03 (1.02, 1.05) |
| **Sensitivity Analysis (restricted to only those patients with a single outpatient visit)** |  |  |  |  |
| ED | 0.86 (0.79, 0.92) | 0.92 (0.88, 0.97) | 0.85 (0.81, 0.88) | 0.93 (0.90, 0.95) |
| Hospitalization | 0.86 (0.76, 0.98) | 0.87 (0.80, 0.94) | 1.12 (1.00, 1.26) | 1.28 (1.20, 1.37) |
| Death | 1.27 (1.04, 1.55) | 1.07 (0.93, 1.22) | 1.90 (1.61, 2.24) | 1.37 (1.22, 1.54) |
| Composite | 0.85 (0.79, 0.91) | 0.91 (0.87, 0.95) | 0.89 (0.86, 0.93) | 0.97 (0.95, 0.99) |
| **Sensitivity Analysis (zero-inflated Poisson regression using first event only)** |  |  |  |  |
| ED | 0.84 (0.81, 0.87) | 0.90 (0.88, 0.91) | 0.89 (0.86, 0.91) | 0.90 (0.89, 0.91) |
| Hospitalization | 0.95 (0.86, 1.05) | 0.96 (0.92, 1.00) | 0.99 (0.87, 1.13) | 0.92 (0.87, 0.98) |
| Composite | 0.97 (0.94, 0.99) | 0.95 (0.94, 0.96) | 0.93 (0.91, 0.95) | 0.91 (0.90, 0.92) |

\*adjusted for age, sex, Pampalon Deprivation Index, and Charlson score

While absolute outcome rates were substantially lower in patients without ACSC, similar patterns (less ED visits but slightly more deaths) were seen as for patients with ACSC (eFigures 3 and 4): total composite events were less after virtual outpatient visits at 30 days (4.5% vs. 4.9%, aOR 0.96 [0.94-0.97]) but not at 90 days (12.3% vs. 12.5%, aOR 1.03 [1.02-1.05]).

Our two sensitivity analyses (the first restricted to patients with only one outpatient visit in each followup period [30 or 90 days] and the second analyzing only the first event per followup period) were very similar to the primary analyses, confirming the robustness of our findings (Table 4).

## DISCUSSION

Our findings that Albertan adults were less likely to visit a physician, present to an ED, or to be hospitalized in the first year of the COVID-19 pandemic are not unexpected given public health messaging early in the pandemic. Several studies in other jurisdictions have also demonstrated that ED visits and hospitalizations for non-COVID-19 diagnoses declined significantly in 2020 compared to prior years.[13-16] Concerningly, other studies also reported excess out-of-hospital deaths during the early phases of the pandemic[17-19] and it is estimated that between one quarter and one half of the excess all-cause deaths in North America during 2020 did not have SARS-CoV-2 infection.[20-24] These patterns are particularly worrisome since inpatient and outpatient visit rates after the 2003 SARS-CoV-1 epidemic did not recover to pre-epidemic levels in affected cities until several years later.[25] Indeed, recent reports suggest that although outpatient visit rates have increased in the US since the almost 60% decline seen in the early weeks of the pandemic, they have still not recovered to pre-pandemic levels.[26,27]

Our finding that outpatient care shifted from an almost exclusively in-person model pre-pandemic to a mixed model with over 41% of all visits being virtual in the first year of the COVID-19 pandemic is also not unexpected and mirrors the findings from earlier studies. However, while those studies focused on selected populations, selected diagnoses, selected visits (primary care only), or only examined data from the first 3-4 months of the pandemic,[1-10] we were able to examine all outpatient visits (primary care and specialty) for any cause in an entire Canadian province over the first year of the pandemic, which included the lulls between COVID-19 waves.

Our findings of differences in short-term outcomes in the subsequent 1 to 3 months after virtual visits compared to in-person visits (significantly and substantially fewer ED visits balanced against slightly higher mortality risk, albeit also statistically significant), both for patients with and without ACSC, highlights that the two types of outpatient visits should not simply be considered interchangeable. Of course, as our study is observational we cannot make determinations of causality. A number of factors which may have influenced physician or patient decisions about type of outpatient follow-up could have biased outcomes. For example, it is possible that virtual visits were done preferentially in sicker, frailer patients or those with more comorbidities due to concern that their risks from potential SARS-CoV-2 exposure were higher than for other patients (so called collider bias) or because they had more difficulty in physically attending clinics. Alternately, it is also plausible that some physicians chose to see patients who were sicker or frailer in-person to see if they could stave off ED visits. Ultimately, to evaluate the impact of virtual care properly requires a randomized trial, which was not possible given the pandemic realities, and ideally a wider spectrum of outcomes, including patient and provider reported experience measures. Regardless, our findings do raise questions about the equivalency of outpatient visit types that warrant further study.

Our finding that some outcomes (medication dispensations and frequency of followup for patients with ACSC) were similar after the rapid shift towards a mixed in-person/virtual model for outpatient care in the first year of the pandemic provides some reassurance about the care of those already receiving treatment for chronic conditions. This is consistent with the demonstration in another Canadian province[1] that outpatient visits declined least among those with the highest health care needs. However, we did not have access to medication dosages prescribed - this is a potentially important limitation since one recent study[9] reported that heart failure patients were 61% less likely to have their antifailure therapy intensified after a virtual visit than an in-person visit and two other studies[7,8] reported that virtual visits were associated with 38% fewer new prescriptions than in-person visits. Thus, further research is needed to investigate whether the shift towards virtual outpatient care has negatively impacted medication intensification for chronic conditions such as heart failure, hypertension, diabetes, or dyslipidemia.

Further research is also needed to investigate whether the shift towards virtual outpatient care has negatively impacted diagnostic test ordering practices, particularly screening for and detection of new conditions (such as diabetes, atrial fibrillation, or cancers). For example, a recent US study[6] revealed that while the total number of outpatient visits (virtual and in-person) only decreased by 21% in the second quarter of 2020, assessments of blood pressure decreased by 50% and cholesterol by 37%. Another US academic health system reported that 6 primary care screening quality measures were done less than one-third of usual frequency in the early months of the pandemic and 4 were still being performed at less than two-thirds of pre-pandemic levels even in the lull between the first and second waves.[28] It is not surprising that reports are now also emerging of declines in new cancer diagnoses during the COVID pandemic.[29]

The impact of virtual visits on continuity of care is also uncertain, an important factor to evaluate since patients who report higher care continuity (based on face-to-face encounters) exhibit greater satisfaction, better quality of care, better medication adherence, fewer ED visits, fewer hospitalizations, and fewer deaths.[30-33]

While this study includes data from an entire population in a universal access, single payer health care system without user fees at the point of care, there are some limitations to our data. For one, the generalizability of our findings to other health care systems are uncertain. For example, while a US study [2] reported substantial variation in the use of virtual visits between regions and socioeconomic classes (with 28% less use by individuals living in socially disadvantaged neighbourhoods) raising concerns about virtual care accentuating healthcare disparities[34], we did not find such a pattern in Alberta. Second, we did not assess reasons for, or content of, visits and therefore could not assess the appropriateness, quality, or cost-effectiveness of virtual visits. This is an open question as one US study of nearly 37 million commercially insured individuals reported that annual healthcare costs were 65% higher in people with at least one virtual visit in 2020 compared to those with only in-person ambulatory visits in 2020.[2] Third, patient ethnicity is not captured in the healthcare datasets we used. Fourth, we do not know whether it was the physician or the patient who decided which visits should be virtual and which in-person. Fifth, we do not know the extent to which changes in the availability of personal protective equipment or patient vaccination status may have altered the balance between virtual and in-person visits (vaccines began to be rolled out in late December 2020 for high risk individuals in Alberta but were not available to the general public until February 2021).

In conclusion, we found that the shifts in the type of outpatient visits caused by the COVID-19 pandemic in the Canadian province of Alberta appeared to maintain existing care and prescribing patterns for patients with ACSC. However, virtual visits were associated with significantly fewer subsequent healthcare encounters compared to in-person visits for patients with and without ACSC. Whether long-term outcomes will also be different as a result of the increase in virtual care is still unknown and certainly a possibility given declines in screening activities and less personalized case management (such as medication intensification) for patients with chronic conditions. There is an urgent need for research to define which patients and which conditions are most suitable for virtual outpatient followup and, as with all outpatient care, the optimal frequency of such visits.

## Supporting information

STROBE checklist

## Data Availability

To comply with Alberta Health Information Act the dataset used for this study cannot be made publicly available. The dataset from this study is held securely in coded form within the AbSPORU (Alberta Support for Patient Oriented Research Unit) Data Platform. While legal data sharing agreements between the investigators, AbSPORU, and Alberta Health Services/Alberta Health prohibit us from making the dataset publicly available, access may be granted to those who meet pre-specified criteria for confidential access, available at www.absporu.ca. The dataset analytic codes are available from the authors upon request, understanding that the computer programs may rely upon coding templates or macros that are unique to AbSPORU.

## Data Availability Statement

To comply with Alberta’s Health Information Act, the dataset used for this study cannot be made publicly available. The dataset from this study is held securely in coded form within the AbSPORU (Alberta Support for Patient Oriented Research Unit) Data Platform. While legal data sharing agreements between the investigators, AbSPORU, and Alberta Health Services/Alberta Health prohibit us from making the dataset publicly available, access may be granted to those who meet pre-specified criteria for confidential access, available at www.absporu.ca. The dataset analytic codes are available from the authors upon request, understanding that the computer programs may rely upon coding templates or macros that are unique to AbSPORU.

## Acknowledgements

This study is based in part on data provided by Alberta Health and Alberta Health Services. The interpretation and conclusions contained herein are those of the researchers and do not represent the views of the Government of Alberta or Alberta Health Services. Neither the Government of Alberta nor Alberta Health Services express any opinion in relation to this study.

---

**eFigure 1:**
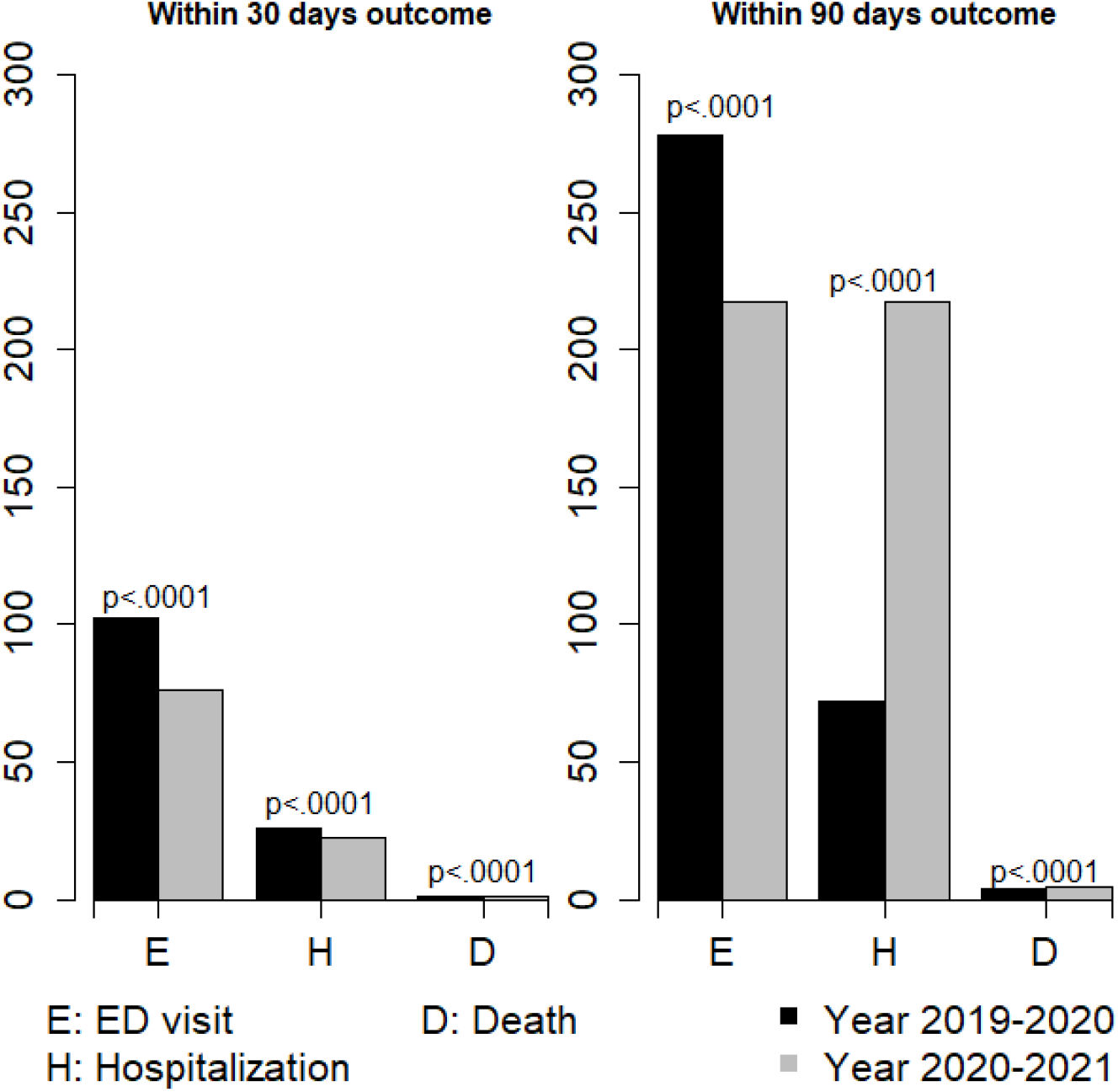
Outcomes within 30 days and 90 days after outpatient physician visits in the year before and the first year of the COVID-19 pandemic for patients with ACSC, reported per thousand visits.

**eFigure 2:**
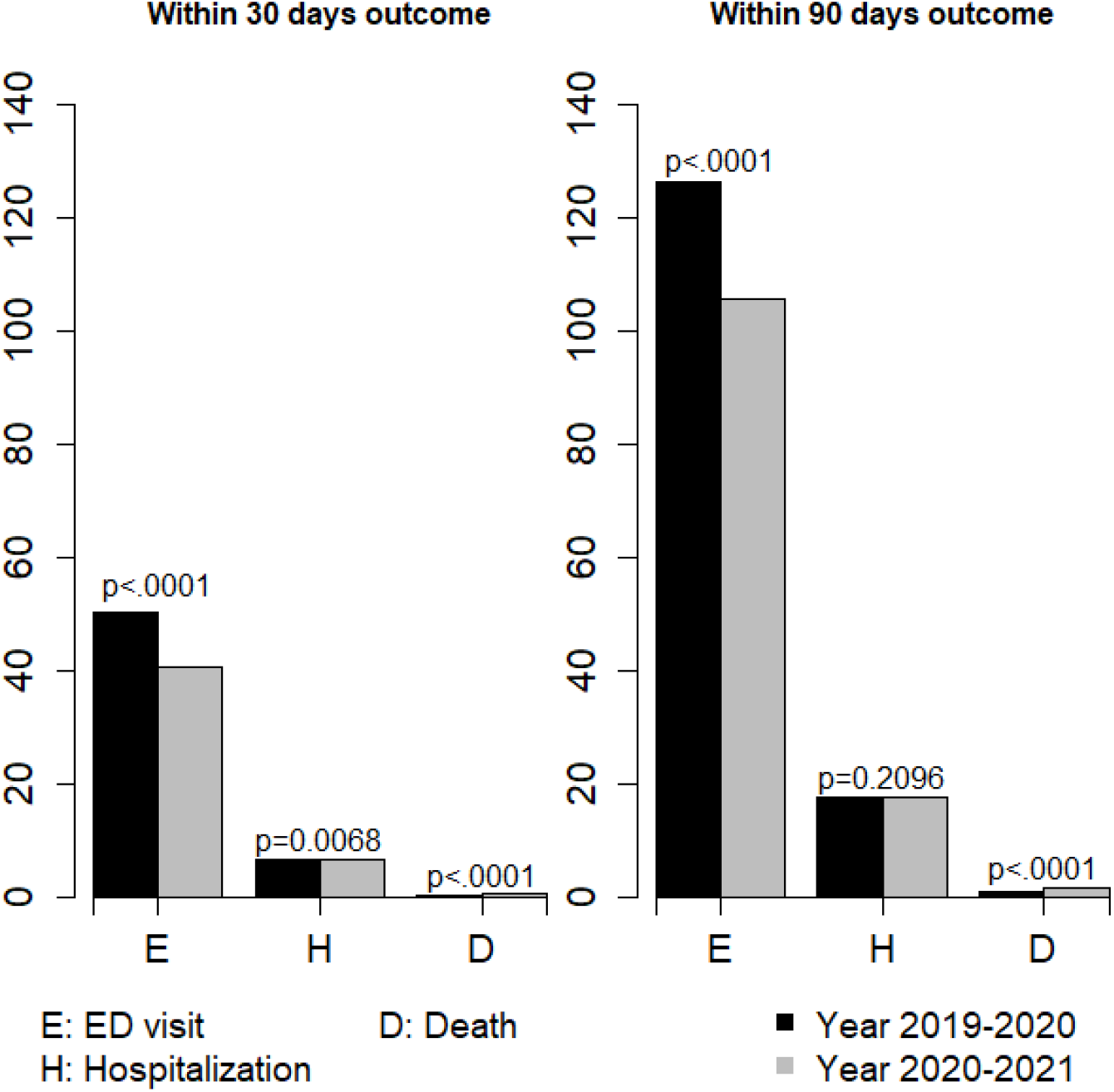
Outcomes within 30 days and 90 days after outpatient physician visits in the year before and the first year of the COVID-19 pandemic for patients without ACSC, reported per thousand visits.

**eFigure 3:**
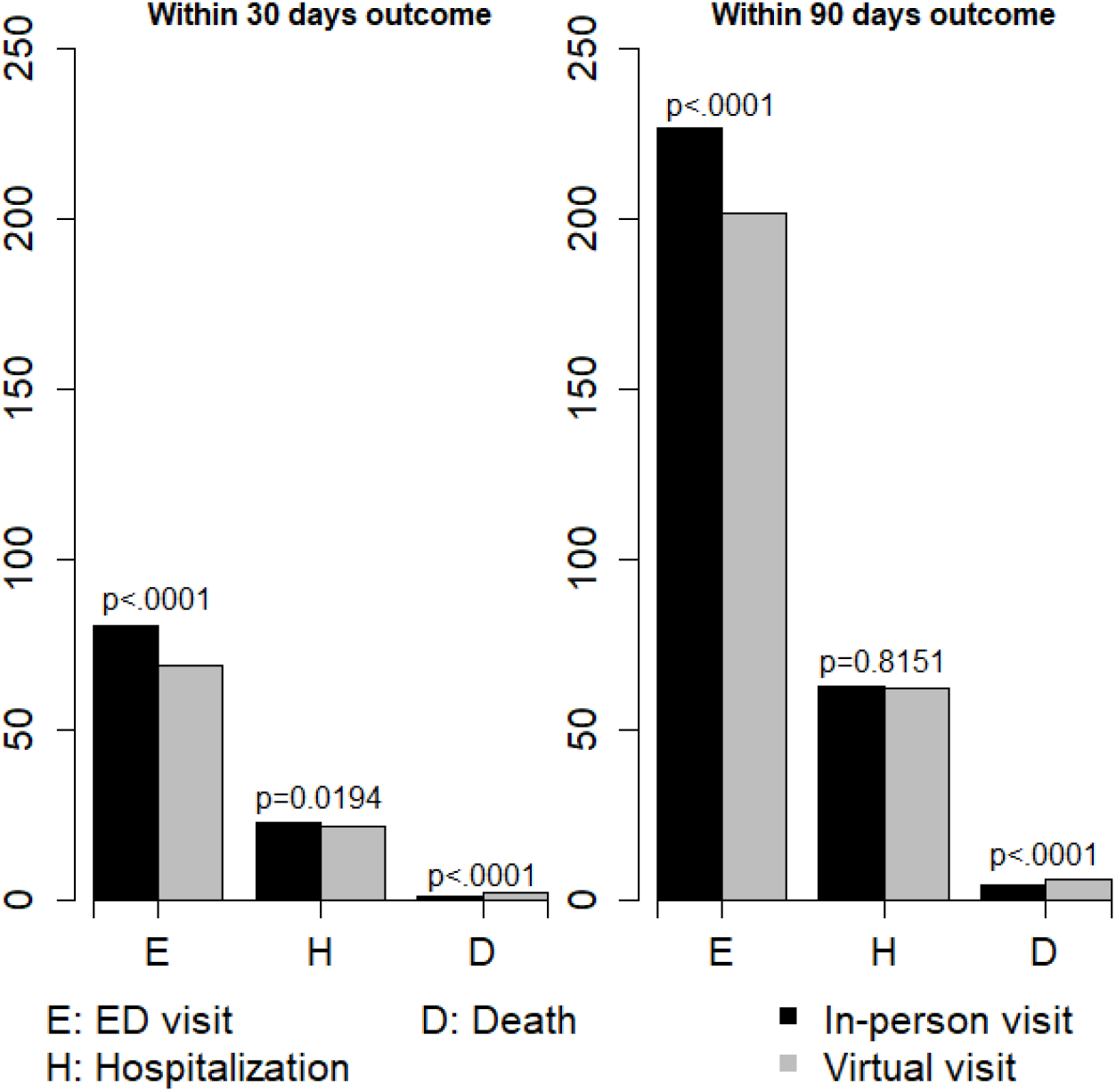
Outcomes within 30 days and 90 days of virtual versus in-person visits for ACSC patients during the first year of the pandemic, reported per 1000 outpatient visits.

**eFigure 4:**
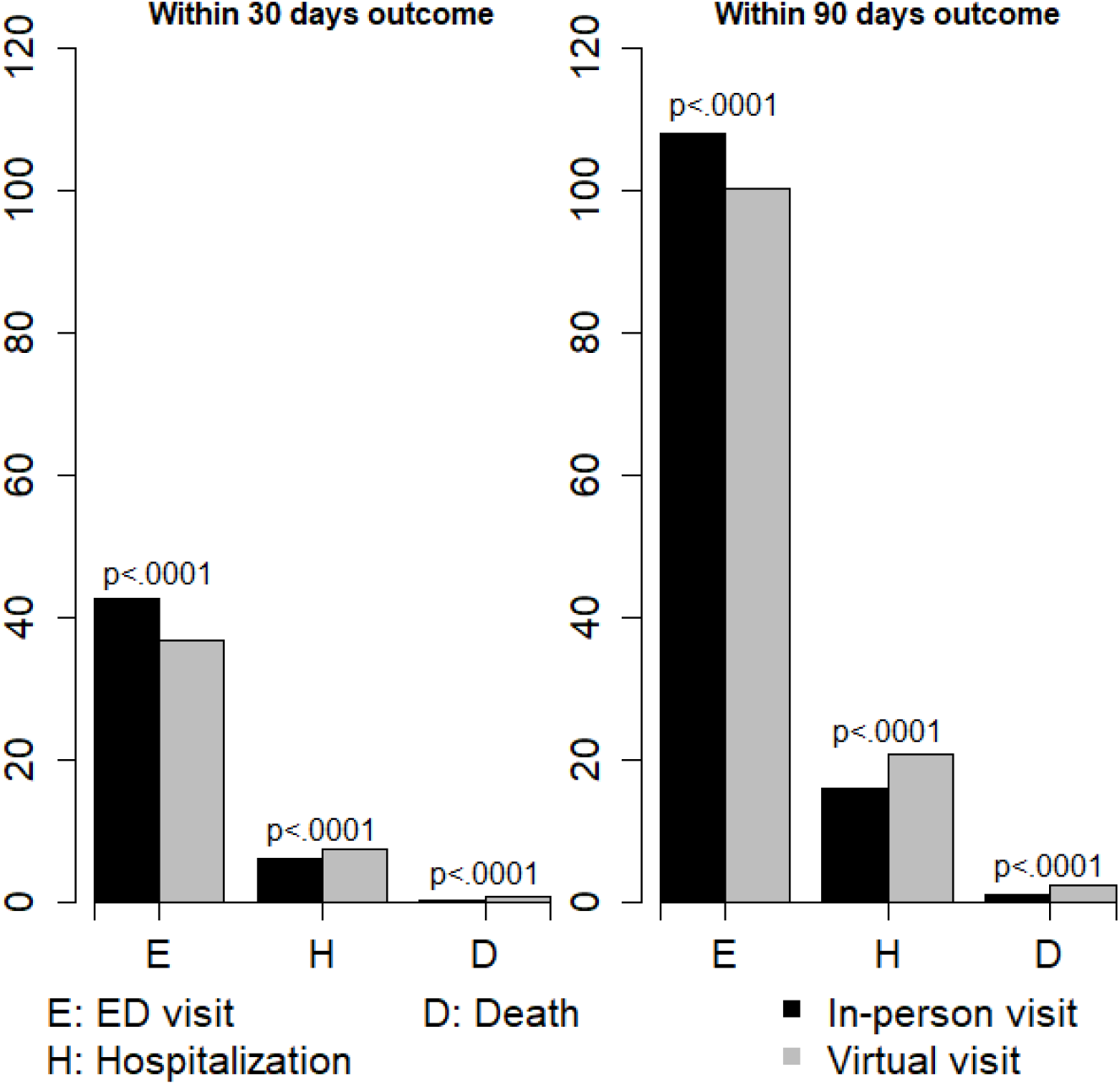
Outcomes within 30 days and 90 days of virtual versus in-person visits for patients without ACSC during the first year of the pandemic, reported per 1000 outpatient visits.

**eAppendix Table 1:**
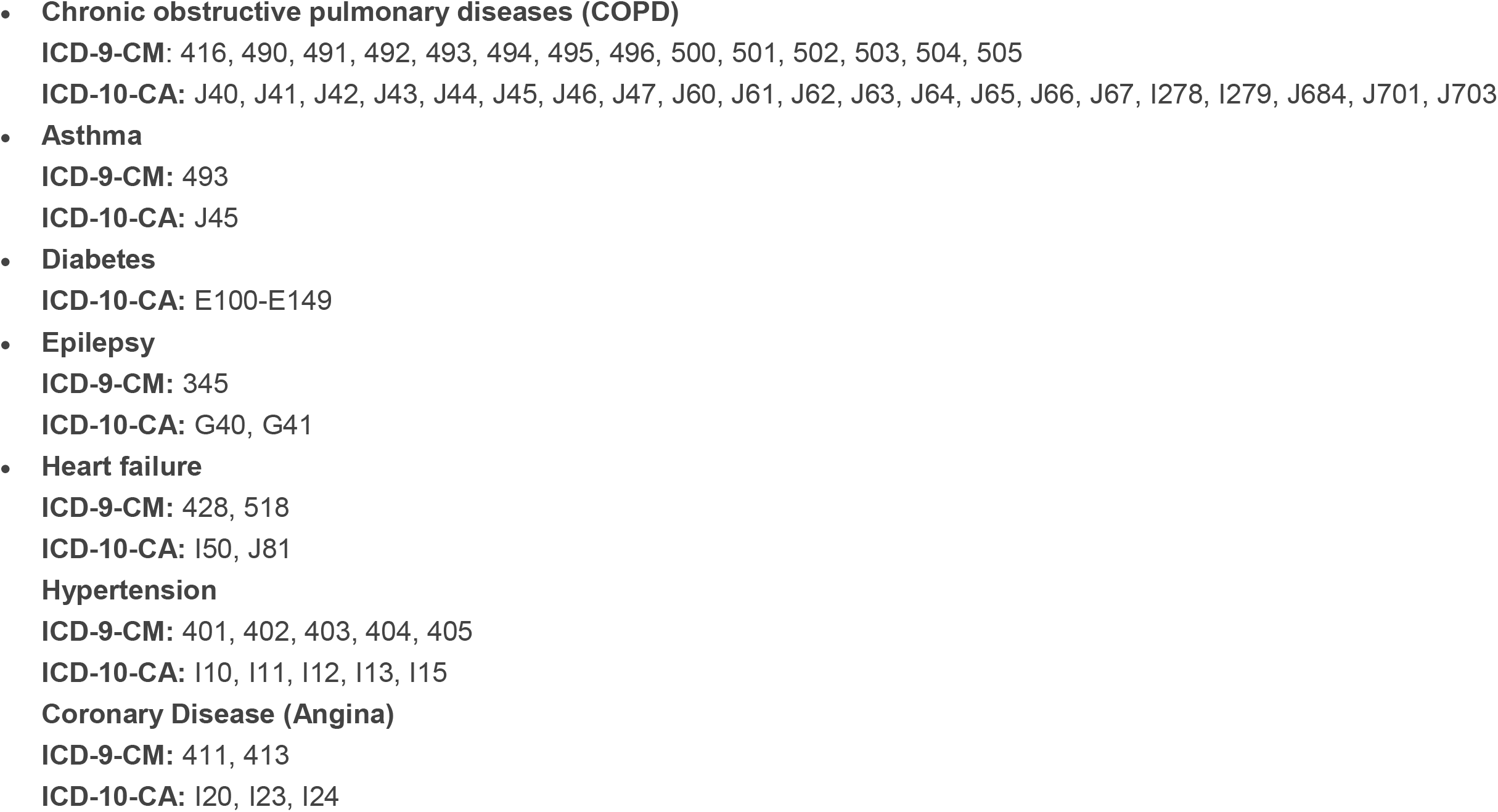
List of case definitions for ambulatory care sensitive conditions. Defined by 1 hospitalization or 1 ED visit or 2 Practitioner Claims in the year of study period.

